# Machine Learning-Driven Assessment of Early Graft Function in Living Donor Kidney Transplantation Using Intraoperative Laser Speckle Contrast Imaging

**DOI:** 10.1101/2025.10.07.25336974

**Authors:** Yitian Fang, Li You, Hendrikus J.A.N. Kimenai, Miao-Ping Chien, Frank J.M.F. Dor, Ron W.F. de Bruin, Robert C. Minnee

**Author notes:** Authors contributed equally (Yitian Fang, Li You).

## Abstract

**Background:** Although living donor kidney transplantation (LDKT) generally achieves excellent outcomes, 5–12% of recipients experience early graft dysfunction, which is associated with poorer long-term survival. Current predictive tools rely mainly on clinical parameters and lack intraoperative applicability. Laser speckle contrast imaging (LSCI) enables real-time, non-contact assessment of renal microcirculation, and may provide complementary insight when integrated with machine learning (ML).

**Methods:** In this prospective cohort study, we performed intraoperative LSCI measurement in 110 adult LDKT recipients at Erasmus Medical Center. Early graft function was assessed by estimated glomerular filtration rate (eGFR) at 1 week posttransplant, with patients classified as Group I (eGFR ≥ 30 mL/min/1.73 m²) and Group II (eGFR < 30 mL/min/1.73 m²). Two predefined feature sets were used for model development: (i) a Clinical Model (selected clinical variables) and (ii) a Combined Model (clinical + convolutional neural network [CNN]-derived LSCI features). Four ML algorithms (support vector machine [SVM], logistic regression, random forest [RF], and XGBoost) were trained using 5-fold cross-validation with Synthetic Minority Oversampling Technique (SMOTE) and evaluated on independent test sets across 30 repeated iterations.

**Results:** Of 110 recipients, 15 (17%) had eGFR < 30 mL/min/1.73 m² at 1 week. Patients in Group II received kidneys from older donors with lower predonation eGFR, had higher BMI, more cardiovascular comorbidity, and greater intraoperative blood loss. The Combined Model consistently outperformed the Clinical Model across all algorithms. For example, SVM achieved higher accuracy (0.89 [95% CI, 0.85–0.92] vs. 0.79 [0.75–0.84]) and logistic regression yielded higher recall (0.86 [0.83–0.88] vs. 0.76 [0.74–0.79]). In independent test sets, Combined Models maintained better performance, with SVM achieving the highest F1 score (0.60 [0.50–0.71]) and RF achieving the highest recall (0.88 [0.50–1.00]). Grad-CAM visualizations confirmed that CNN-extracted features localized to physiologically relevant perfusion regions. LSCI also enabled real-time detection and correction of vascular complications in two cases.

**Conclusions:** Integrating intraoperative LSCI features with clinical variables using ML significantly improved prediction of low one-week eGFR (< 30 mL/min/1.73m²) in LDKT compared with clinical data alone. LSCI also enabled real-time detection of vascular complications, underscoring its role as both a predictive and intraoperative guidance tool. Larger multicenter studies are warranted to validate its generalizability and explore applications in other transplant scenarios.

## INTRODUCTION

Living donor kidney transplantation (LDKT) provides superior graft survival compared to deceased donor kidney transplantation.^1^ However, a subset of 5-12% LDKT recipients develop early graft dysfunction, which may negatively impact long-term outcome if not promptly identified.^2^

Early graft function is influenced by a complex interplay of donor, recipient, and surgical factors. Previous studies have investigated a range of clinical factors, including demographic characteristics, biological parameters (including kidney allograft function, proteinuria, and circulating anti-HLA antibody specificities and concentrations), and allograft pathology information to identify patients at risk of early graft loss.^3, 4^ While these clinical variables are informative, their predictive accuracy remains limited, particularly in the intraoperative setting where real-time risk assessment could guide immediate clinical decision-making and indicate graft function.

Laser speckle contrast imaging (LSCI) is an emerging optical technique that enables non-invasive, non-contact, and contrast-free visualization of tissue microcirculation in real-time. By capturing dynamic speckle patterns generated by moving red blood cells, LSCI offers a semi-quantitative measure of renal cortical microcirculation. Due to the strong correlation between renal cortical microcirculation and kidney function, LSCI holds valuable functional insight into graft function beyond macroscopic inspection.^5^

With the rise of artificial intelligence in medicine, machine learning (ML) and deep learning approaches have demonstrated substantial potential in supporting clinical decision-making. In particular, convolutional neural networks (CNNs) are a form of artificial neural networks that are able to extract and interpret high-dimensional features, which makes CNN ideal for image analysis.^6^ When combined with traditional clinical prediction models, these CNN-derived imaging features may improve the performance of predictive models.

In this study, we aimed to investigate the value of intraoperative LSCI-derived imaging features, extracted using a CNN-based approach, in predicting early graft function following LDKT. We hypothesized that prediction models incorporating both clinical and LSCI data would outperform models based solely on clinical variables, thereby providing a more accurate tool for identifying early risk in LDKT recipients.

## METHODS

### Study Design

We used the Transparent Reporting of a multivariable prediction model for Individual Prognosis Or Diagnosis (TRIPOD) for the reporting of our study (**Supplementary Methods S1**).^7^

A schematic overview of the study design is shown in **Figure 1**. This prospective, single-center cohort study enrolled 110 adult patients who underwent living donor kidney transplantation (LDKT) at Erasmus Medical Center between March 2023 and March 2025. Intraoperative LSCI measurements were performed after reperfusion during the transplant procedure.

**Figure 1.**
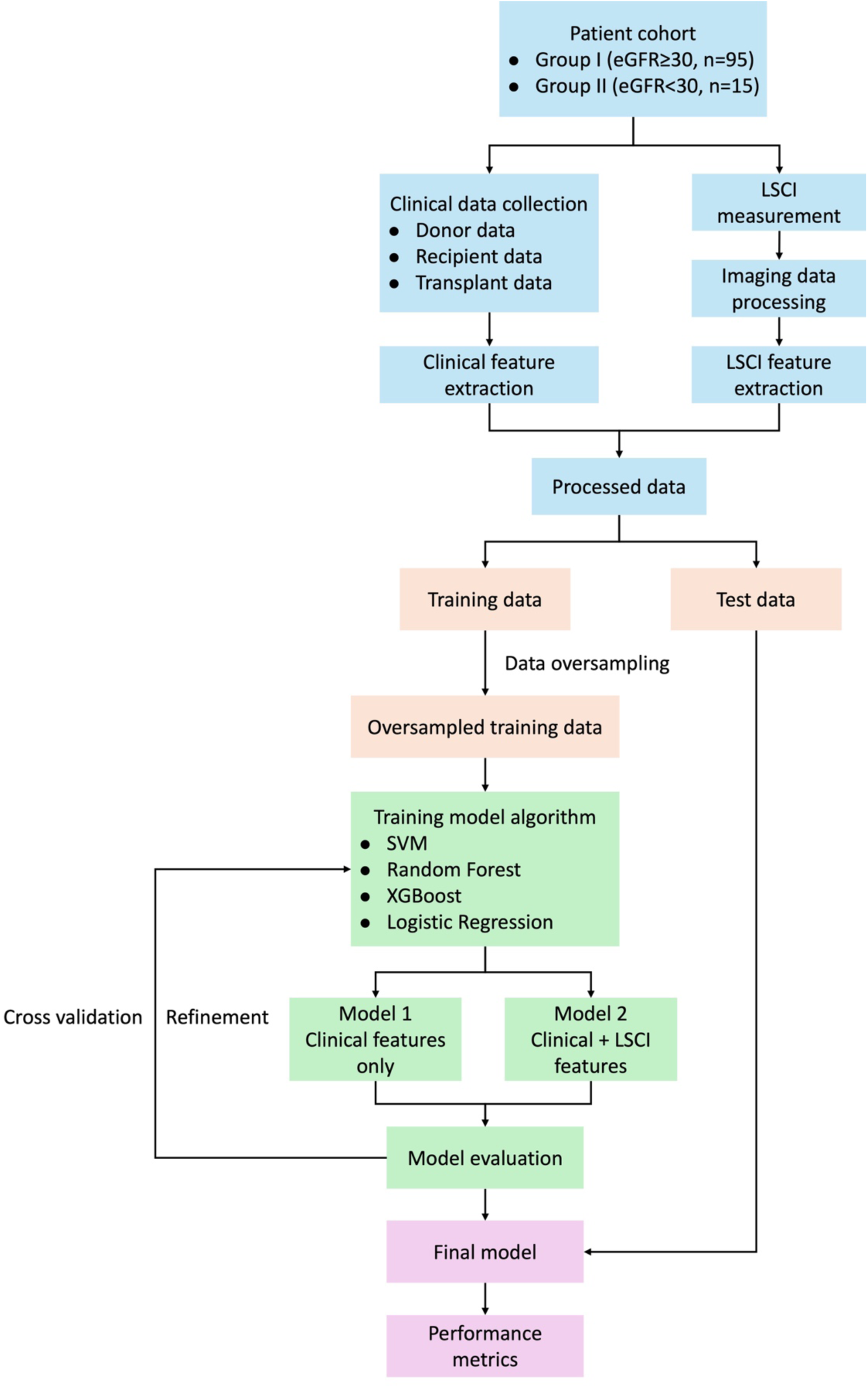
Flowchart for predicting early graft dysfunction in living donor kidney transplantation.

Estimated glomerular filtration rate (eGFR) was assessed one week posttransplant. Patients were classified into two groups based on early graft function. Patients with eGFR ≥ 30 mL/min/1.73m^2^ were classified as Group I, whereas those with eGFR < 30 mL/min/1.73m^2^ were classified as Group II. We selected the threshold of 30 mL/min/1.73 m^2^ because an eGFR below this level is considered as the indication of severe renal impairment and kidney replacement therapy.^8, 9^

To assess the predictive value of intraoperative LSCI, we developed and compared two ML models with predefined feature sets:

(1) Clinical Model, which includes selected clinical variables only; and (2) Combined Model, which includes the same clinical features and CNN-derived LSCI features. Both ML models were trained and evaluated separately, and model performance was assessed using standard classification metrics.

### Clinical Data Collection

To investigate the key prognostic determinants of early graft dysfunction, we included donor-related characteristics, recipient-related characteristics, and transplant data. The following parameters were collected for the construction of the early graft function prediction model: (1) donor age, sex, BMI, and predonation renal function; (2) recipient age, sex, BMI, cause of ESRD, and comorbidities; (3) transplant data including ABO compatibility, virtual panel reactive antibodies (vPRA), HLA-A/B/DR mismatch numbers, preemptive transplant status, warm ischemia time, and estimated intraoperative blood loss.

### LSCI Measurement

LSCI was performed using the MoorO2Flo system (Moor Instruments Ltd., Axminster, UK) to assess renal microcirculation during the early reperfusion phase. All measurements were conducted after reperfusion with the kidney graft positioned in its final transplant orientation. Each recording lasted 10 seconds. To minimize ambient light interference, surgical lights were turned off and external light sources were blocked during measurement.

During data acquisition, real-time LSCI images were monitored by the surgical team. If abnormal perfusion patterns (e.g., focal hypoperfusion) were observed, transplant surgeons were informed to evaluate potential issues such as anastomotic stenosis, renal artery kinking or venous outflow problems. In such cases, a repeat LSCI acquisition was performed following intervention and included for analysis. Detailed working principle, protocol and quality control procedures of the LSCI measurements are described in **Supplementary Methods S2**.

### LSCI Data Processing

The workflow of LSCI measurement, image processing, and feature extraction is illustrated in Figure 2.

**Figure 2.**
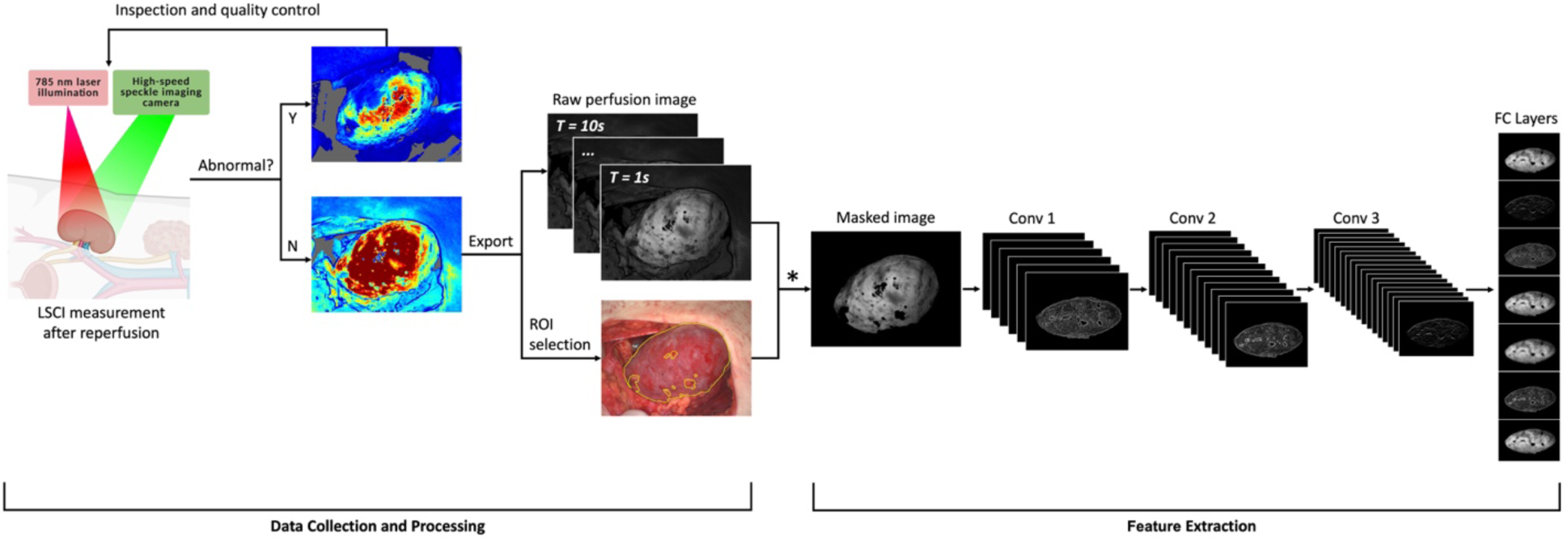
Workflow of LSCI measurement, image processing and feature extraction. LSCI was performed after reperfusion during kidney transplantation. If perfusion abnormalities were observed when real-time monitoring, repeat measurements were performed after corrective intervention and included in the analysis. For each patient, ten representative LSCI frames (one per second) were selected, and both raw perfusion and corresponding color images were exported. Using the color image as anatomical reference, the visible renal surface was manually delineated as the region of interest (ROI), excluding areas obscured by adipose tissue or surgical artifacts. This ROI was applied to the perfusion images to create masked images. The masked images were processed using a customized convolutional neural network (CNN) based on AlexNet architecture, consisting of three convolutional layers followed by two fully connected (FC) layers. The CNN extracted high-dimensional feature vectors from the images, which were subsequently used for predictive modeling.

For each patient, LSCI measurement was performed at a frame rate of 20 Hz over a duration of 10 seconds, resulting in a total of 201 frames per recording. For data analysis, 10 representative frames were selected-one from each second, corresponding to frames 20, 40, 60, …, 200. From each selected frame, both the LSCI perfusion image and the corresponding colour image were exported.

Due to the limited measurement depth of LSCI (∼0.3 mm), surface obstructions can interfere with accurate perfusion measurements.^10, 11^ Using the color images as anatomical reference, the entire visible kidney surface was manually delineated as the region of interest (ROI), excluding areas obscured by adipose tissue or surgical artifacts such as cauterized clots. The defined ROI for each kidney was applied to the corresponding LSCI perfusion image to create a masked image.

### LSCI Image Feature Extraction

The masked perfusion images were used as input for our CNN, which in this study, is based on a modified AlexNet architecture consisting 3 convolutional layers followed by 2 fully connected layers.^12^ Convolutional layers are used to iteratively extract LSCI image features, such as ROI shapes, textures, brightness, brightness gradient, colormaps, or abstract patterns that can describe other spatial structures. Let *f* = {*f*_1_, *f*_2_, …, *f_n_*} denote the sets of *n* features. Fully connected layers will make use of all extracted features and classify whether an image represents “Group I” (represented by 0 in a binary case) or “Group II” (represented by 1 in a binary case). The classification is done via a loss function ℒ(*f*) → 1.

The supervised learning process is summarized as follows: (1) Each input image has a ground truth label. Let *I* = {*I*_1_, *I*_2_, …, *I_m_*} denote the set of *m* input images, and *L* = {*L*_1_, *L*_2_, …, *L_m_*} the set of corresponding ground truth labels, where *L*_i∈[1,_*_m_*_]_ = 0 *or* 1. (2) CNN extracts a set of features *f* = {*f*_1_, *f*_2_, …, *f_n_*} from *I* and assigns weights *w* = {*w*_1_, *w*_2_, …, *w_n_*} to them. Initially, the weights are equal, and through learning the network adjusts these weights so that certain features become more important in predicting the output. (3) The weighted features are passed through a loss function to calculate a predicted label ℒ(*f*, *w*). The loss function quantifies the difference between the predicted and ground truth labels and updates the weights accordingly. A full pass through all training images constitutes one epoch. In our set up, we trained the CNN for 50 epochs, meaning CNN scanning through all images *I* and update weights *w* for 50 times, which is a standard number of epochs to achieve stable performance.

Through supervised learning with AlexNet, we developed a CNN model that achieved a classification accuracy of 82%. This process yielded both features and their corresponding weights, which reflect their relative importance in distinguishing between outcome classes. Features with positive weights (*w_i_* > 0) were considered informative and selected for further analysis.

### LSCI Image Feature Localization

The selected CNN features can be pin-pointed back to the original LSCI data for sanity check. **Figure 3** illustrates feature localization using Gradient-weighted Class Activation Mapping (Grad-CAM) across two convolutional layers of the CNN trained on LSCI data. Grad-CAM generates class-specific heatmaps by backpropagating the gradients of the predicted class to the convolutional feature maps, thereby highlighting regions most influential for the network’s decision.^13^

**Figure 3.**
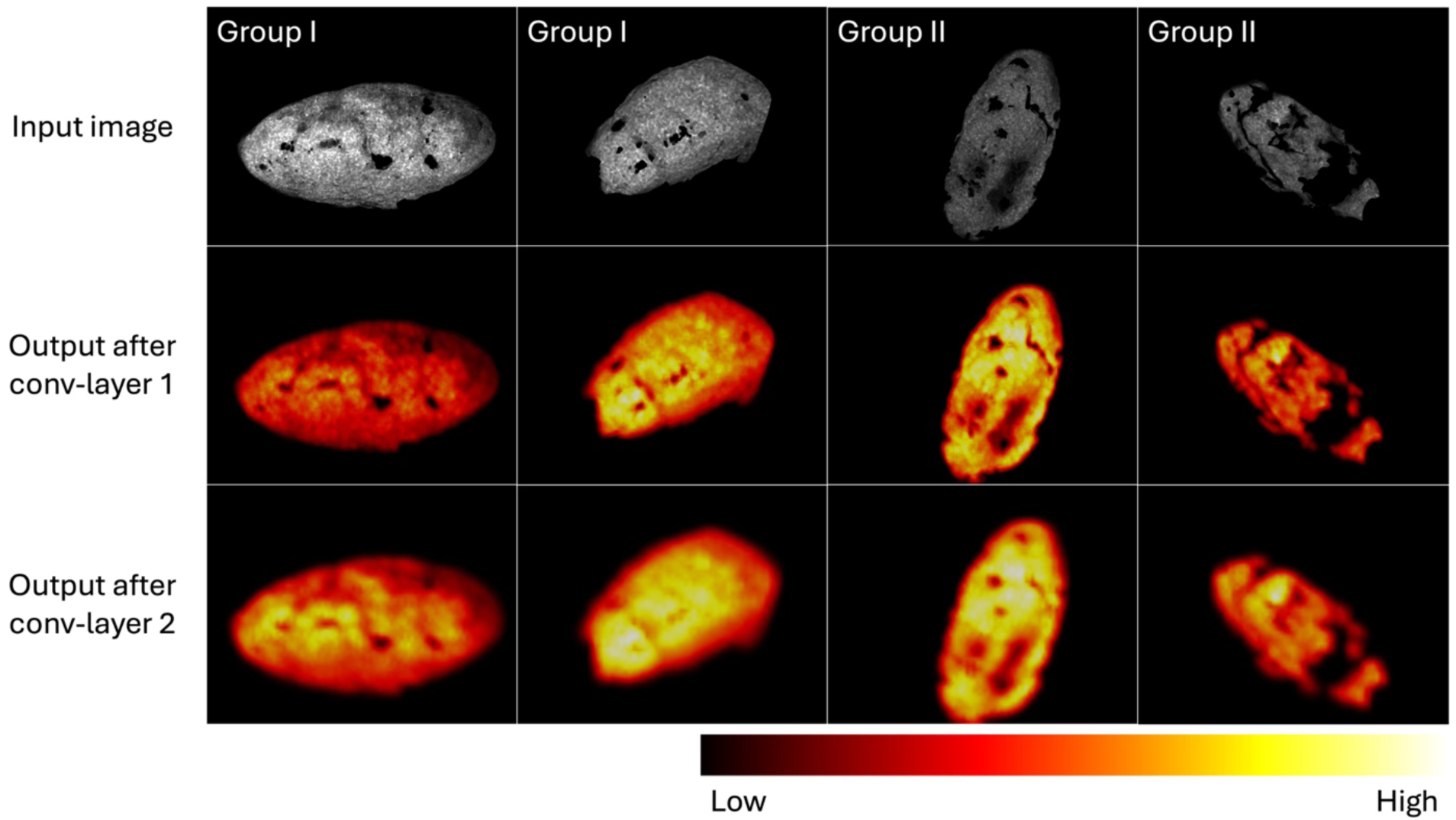
Visualizations of convolutional feature activations from the CNN trained on LSCI data acquired during kidney transplantation. The top row shows representative raw LSCI input images, labelled as either Group I or Group II. The second and third rows illustrate the Grad-CAM heatmaps from the first and second convolutional layers, respectively. The first layer activations are broadly distributed, reflecting low-level texture and contrast patterns, whereas the second layer activations become more refined and spatially focused. The highlighted regions (e.g., yellow region) indicate areas most influential in the network’s decision.

The first row shows representative raw LSCI input images, categorized as stable or at-risk grafts. The second and third rows display the corresponding Grad-CAM heatmaps from the first and second convolutional layers, respectively. In the first layer, activations are distributed broadly and capture low-level texture and contrast variations. In contrast, the second layer produces more refined and spatially focused activations, highlighting regions most discriminative for graft function classification. These visualizations indicate that the CNN focused on physiologically relevant regions when making predictions.

### Statistical Analysis

Continuous variables were described using medians with interquartile ranges (IQR), and categorical variables were described using number and percentages. Medians and proportions between groups were compared with the Mann–Whitney U test, or the *x*^2^ test (or Fisher exact test if appropriate). Values of *P* < 0.05 were considered significant, and all tests were 2 tailed.

### ML Model Development

We implemented ML models that can be applied to classify binary outcomes, referred to as ML classification models, including Support Vector Machine (SVM) with a linear kernel, Extreme Gradient Boosting (XGBoost), Random Forest, and Logistic Regression. Each ML model is detailed in the **Supplementary Methods S3**.

All predictor variables were centered and scaled. The full cohort was randomly split into training (70%) and test (30%) sets using stratified sampling. The test set (30%) serves as unseen data to the trained model, used to examine the robustness of model training.

For training set, the Synthetic Minority Oversampling Technique (SMOTE) was applied to address class imbalance before model fitting. Predictor variables were selected based on the concordance index (C-index) of each variable with respect to the outcome using original dataset, which quantifies the variable discriminatory ability. Variables with a C-index > 0.6 or < 0.4 were included in subsequent model development. Model training was conducted on the SMOTE-balanced training dataset. ML was performed using 5-fold cross-validation with a grid search strategy, with 4 folds used to train the model and the left-out fold used to evaluate the model’s performance.

### Model Evaluation

Each model was trained and evaluated over 30 independent iterations with different random stratified splits to ensure performance robustness. In each iteration, the best-performing parameter combination was identified based on cross-validated AUC within the SMOTE-balanced training set. The final tuned model was then evaluated on the held-out test data for external validation. Performance metrics (accuracy, precision, recall, and F1 score) were calculated separately for the training and test datasets and summarized across iterations.

## RESULTS

### Participant Characteristics

A total of 110 LDKT patients were enrolled in the study, including 95 (83%) cases in Group I and 15 (17%) in Group II.

Baseline characteristics stratified by graft function are displayed in **Table 1**. Compared to Group I, patients in Group II received kidneys from older donors (median 64 [IQR 56-68] vs. 55 (47-64) years; p=0.027) with lower donor pre-donation eGFR (74 [70-83] vs. 87 [76-97] ml/min/1.73m^2^; p=0.011). Patients in Group II had significantly higher BMI (29.2 [25.6-32.7] vs. 25.5 [23.5-28.0] kg/m^2^; p=0.022), and were more likely to have cardiovascular disease (60.0% vs. 26.3%; p=0.015), and higher intraoperative blood loss (250 [200-375] vs. 100 [100-200] mL; p<0.001).

**Table 1.** Cohort’s characteristics.

|  | **Group I**<br>**(n=95)** | **Group II**<br>**(n=15)** | **P value** |
| --- | --- | --- | --- |
| **Donor characteristics** |  |  |  |
| Age, yr, median (IQR) | 55 (47-64) | 64 (56-68) | 0.027 |
| Male sex, n (%) | 42 (42.2) | 7 (46.7) | 1.000 |
| BMI, median (IQR) | 26.1 (23.8-28.1) | 26.5 (23.4-28.0) | 0.764 |
| Serum creatinine, $\mu\text{mol/l}$, median (IQR) | 76 (69-87) | 80 (71-92) | 0.278 |
| eGFR, ml/min per 1.73m<sup>2</sup>, median (IQR) | 87 (76-97) | 74 (70-83) | 0.011 |
| **Recipient characteristics** |  |  |  |
| Age, yr, median (IQR) | 56 (47-66) | 63 (53-69) | 0.190 |
| Male sex, n (%) | 59 (62.1) | 7 (46.7) | 0.395 |
| BMI, median (IQR) | 25.5 (23.5-28.0) | 29.2 (25.6-32.7) | 0.022 |
| End-stage renal disease causes, n (%) |  |  | 0.274 |
| Glomerular disease | 24 (25.3) | 2 (13.3) |  |
| Vascular disease | 20 (21.1) | 6 (40.0) |  |
| Diabetes | 12 (12.6) | 3 (20.0) |  |
| Others | 39 (41.1) | 4 (26.7) |  |
| Comorbidities, n (%) |  |  |  |
| Cardiovascular disease | 25 (26.3) | 9 (60.0) | 0.015 |
| Diabetes | 18 (18.9) | 5 (33.3) | 0.302 |
| Hypertension | 49 (51.6) | 7 (46.7) | 0.940 |
| **Transplant characteristics** |  |  |  |
| ABO-incompatible, n (%) | 9 (9.5) | 1 (6.7) | 1.000 |
| Preemptive, n (%) | 58 (61.1) | 8 (53.3) | 0.777 |
| Warm ischemia time, min, median (IQR) | 18 (16-22) | 20 (17-25) | 0.256 |
| Blood loss, mL, median (IQR) | 100 (100-200) | 250 (200-375) | <0.001 |
| **Immunologic parameters** |  |  |  |
| vPRA (%), median (IQR) | 0 (0-24.3) | 0 (0-2.5) | 0.548 |
| HLA-A/B mismatch number, median (IQR) | 2 (2-3) | 3 (2-3.5) | 0.387 |
| HLA-DR mismatch number, median (IQR) | 1 (1-2) | 1 (1-2) | 0.394 |

BMI, body mass index; eGFR, estimated glomerular filtration rate; HLA, human leukocyte antigen; vPRA, virtual panel reactive antibody.

### Intraoperative Detection of Vascular Complications

In our cohort, two cases of perfusion abnormalities were identified intraoperatively using LSCI. The first case demonstrated extensive cortical hypoperfusion caused by anastomotic stenosis following arterial reconstruction due to dual renal artery branches. The anastomosis was removed and reconstructed, after which repeat LSCI confirmed restoration of perfusion (**Figure S1A**). The second case showed localized perfusion deficiency after reperfusion, which was attributed to renal artery kinking. Adjustment of the graft position corrected the defect, and subsequent LSCI imaging confirmed satisfactory cortical perfusion (**Figure S1B**).

### Variable Selection for Model Development

The C-index was calculated for each candidate variable to assess the individual predictive capability (**Figure S2, S3**). Variables with a C-index > 0.6 or < 0.4 were selected for model development, indicating strong discriminatory power in either direction.

For the Clinical Model, the following variables were selected for ML model development: donor age, donor pre-donation eGFR, recipient age, BMI, history of cardiovascular disease, and estimated intraoperative blood loss. For the Combined Model, these six clinical variables were supplemented with five LSCI-derived features to incorporate quantitative perfusion information (**Figure 4**).

**Figure 4.**
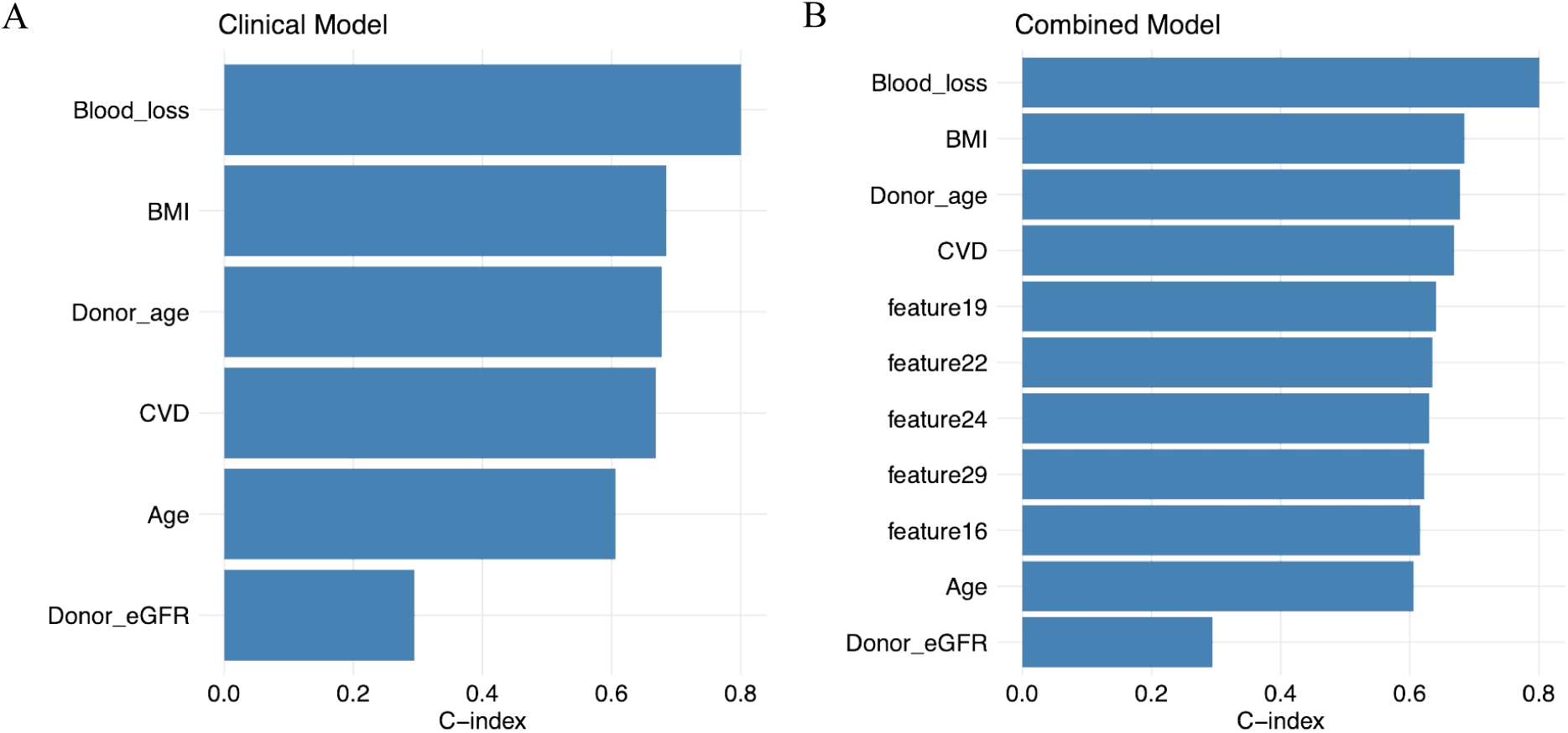
Variables selected for machine learning model development based on concordance index (C-index). (A) Variables selected for the Clinical Model and (B) for the Combined Model. Features with a C-index > 0.6 or < 0.4 were retained for model development. The Combined Model includes the same clinical variables as the Clinical Model along with additional LSCI-derived features.

### ML Model Performance

Model performance was assessed using classification metrics as shown in **Figure 5**. Across all metrics (accuracy, precision, recall, and F1 score), the Combined Model consistently outperformed the Clinical Model during training, regardless of the ML algorithm used. For example, SVM achieved higher accuracy with combined features (0.89 [0.85-0.92] vs. 0.79 [0.75-0.84]; **Figure 5A**, left), while LR yielded a higher recall in the Combined Model (0.86 [0.83-0.88] vs. 0.76 [0.74-0.79], **Figure 5C**, left).

**Figure 5.**
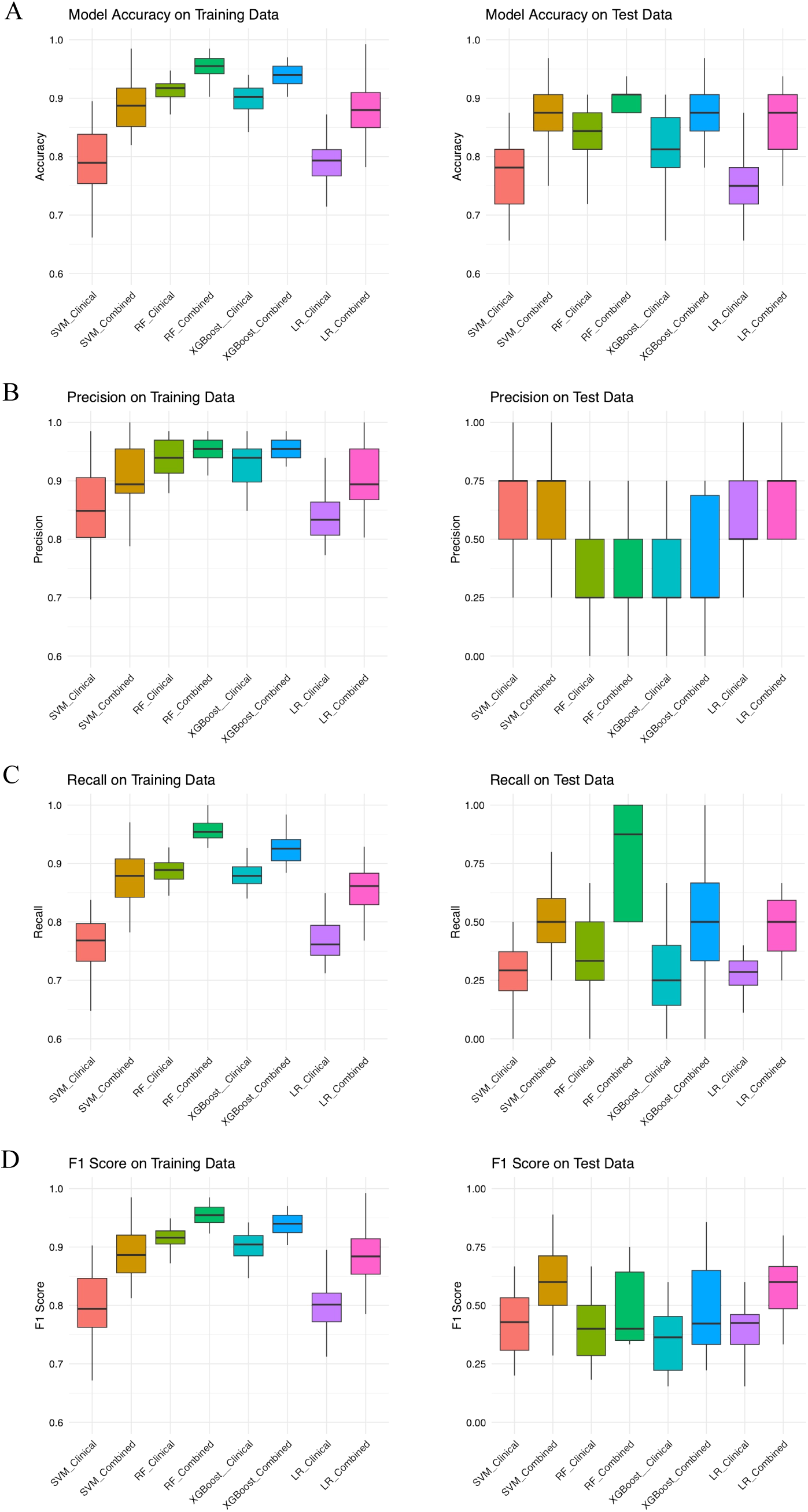
Prediction performance of classification models on SMOTE-balanced training and independent test data. Boxplots show the distribution of (A) accuracy, (B) precision, (C) recall, and (D) F1 score for eight classification models across 30 repeated stratified train/test splits. Each model was trained and cross-validated using SMOTE-balanced training data (left) and tested in an independent test data (right). Models include support vector machine (SVM), random forest (RF), extreme gradient boosting (XGBoost), and logistic regression (LR), each developed using either clinical variables only (Clinical) or combined clinical and LSCI features (Combined).

The prominent advantage of the Combined Model remained when applied on the unseen test dataset. All Combined Models achieved comparable accuracy (**Figure 5A**, right). SVM and LR achieved the highest precision, indicating stronger performance in identifying ‘at-risk’ cases (0.75 [0.50-0.75]; **Figure 5B**, right). In contrast, RF exhibited the highest recall, suggesting greater sensitivity in detecting at-risk cases (0.88 [0.50-1.00] vs. 0.33 [0.25-0.50], **Figure 5C**, right). Among all models, SVM achieved the highest F1-score on the test set, demonstrating the best balance between precision and recall (0.60 [0.50-0.71], **Figure 5D**, right). Full performance metrics are provided in **Table S1** and **S2**.

## DISCUSSION

In this prospective study, we investigated the added value of intraoperative LSCI features, extracted using CNN, for predicting early graft dysfunction following LDKT. Our findings demonstrate that incorporating CNN-derived perfusion features from LSCI into ML models significantly improves predictive performance compared to using clinical variables alone.

Patients at risk of early graft dysfunction were characterized by older donor age, reduced donor pre-donation eGFR, higher recipient BMI, increased prevalence of cardiovascular disease, and more intraoperative blood loss. These clinical variables were selected for model development based on their concordance with graft outcomes. When combined with LSCI-derived features, all ML models showed improved accuracy, precision, recall, and F1 score, both in training and independent test sets. Notably, the Combined Model achieved higher sensitivity in identifying at-risk grafts without compromising specificity, suggesting that LSCI-derived features provide meaningful, complementary insight into graft function recovery.

Previous studies have explored various biomarkers, either pretransplant in the donor, or posttransplant in the recipient, for their prognostic relevance. For example, elevated levels of donor-derived plasma mitochondrial DNA and urinary anaphylatoxin C5a have been associated with an increased risk of DGF.^14, 15^ Similarly, biomarkers found in preservation fluid, such as high concentrations of cell-free miR-505-3p and pi isoenzymes of glutathione S-transferase, have shown significant associations with posttransplant graft dysfunction.^16, 17^ Posttransplant biopsies have also offered insights into graft viability. The presence of partial endothelial to mesenchymal transition and strong expression of TGF-β1 in phenotypic changed endothelial cells have been associated poor graft function recovery.^18^ However, these biomarkers typically represent isolated pathways and may not reflect the complete phenotype of the graft.

In parallel, predictive models leveraging large-scale clinical datasets have gained traction.^19–22^ For instance, the iBOX prognostication system integrates functional, immunological, and histological parameters to predict long-term graft outcomes, and is being adopted in clinical practice.^3, 23^ However, it is important to note that such models are specifically designed for predicting long-term outcomes and are not tailored to assess early graft function in the immediate posttransplant period. Other tools such as the Kidney Donor Risk Index (KDRI) and Kidney Donor Profile Index (KDPI) primarily focus on donor-related factors and are intended to estimate the overall quality of the donor organ prior to transplantation.^24–26^ While valuable for organ allocation and long-term risk stratification, these tools lack the ability to capture intraoperative dynamics that may critically impact graft function.

Therefore, complementary strategies, such as intraoperative imaging and machine learning-based integration of perfusion and clinical data, are needed to enhance early risk stratification. Various imaging modalities, such as Doppler ultrasound, indocyanine green fluorescence angiography, photoacoustic imaging, and MRI, have been investigated to assess graft function in transplantation or machine perfusion settings.^27–31^ However, these techniques are either operator-dependent or require the use of contrast agents. In contrast, LSCI is non-contact, contrast-free, and real-time, making it well suited for surgical workflows. Importantly, the developed tool is practical to apply in the clinical setting. Donor and recipient characteristics are routinely collected before transplantation, and LSCI acquisition requires only 10 seconds of recording under standardized conditions. Once LSCI images are processed by the pre-trained CNN model, feature extraction and risk prediction can be performed within seconds. Therefore, for a new patient, all required inputs can be rapidly obtained and integrated into the ML model, allowing near real-time prediction of early graft function without delaying decision-making.

To our knowledge, this is the first study to apply intraoperative LSCI and ML classification on early graft function prediction in kidney transplantation. The use of CNN enabled the extraction of high-dimensional, abstract image features that represent spatial perfusion patterns, allowing ML algorithms to capture complex associations between intraoperative microcirculation and graft function. These CNN-derived features provided a nuanced characterization of microvascular heterogeneity, likely reflecting subtle hemodynamic abnormalities that precede clinically measurable dysfunction. To enhance interpretability, Grad-CAM was used to map class-specific activations back onto the original LSCI images, thereby identifying the most influential regions for classification. This confirmed that the CNN focused on physiologically relevant areas of the graft, differentiating stable from at-risk cases. Together, the integration of LSCI, CNN feature learning, and Grad-CAM visualization provides a novel and interpretable framework for intraoperative graft function assessment in kidney transplantation.

Moreover, our study highlights the added clinical utility of LSCI in detecting intraoperative vascular complications. In two cases, abnormal perfusion patterns identified by LSCI revealed anastomotic stenosis and arterial kinking, both of which were promptly corrected, with restoration of cortical perfusion confirmed by repeat imaging. These findings highlight the ability of LSCI to provide immediate intraoperative feedback, facilitating timely surgical intervention and potentially reducing postoperative complications.

Despite these promising strengths, several limitations should be acknowledged. First, the sample size was limited in the At-risk group, although we employed SMOTE to mitigate class imbalance in the training phase and performed repeated validations to ensure robustness. Second, LSCI measurements, while standardized in this study, may be affected by variations in kidney exposure, lighting conditions, or technical factors. Although we implemented strict quality control protocols, broader generalizability requires validation across institutions. Third, while CNNs offer powerful feature extraction capabilities, the interpretability of these features remains limited. Lastly, the use of estimated GFR at one week posttransplant as a surrogate for early graft function is practical and commonly adopted, but it may not fully capture long-term graft outcomes. Follow-up studies linking early perfusion profiles with long-term graft survival would further validate the clinical relevance of LSCI-guided risk stratification. Also, follow-up studies should also explore the value of LSCI in deceased donor kidney transplantation as it has higher incidence of early graft dysfunction or delayed graft function compared to the living ones.

In conclusion, our findings suggest that CNN-extracted LSCI features, when integrated with clinical variables and analyzed using ML models, significantly improves the prediction of low one-week eGFR (< 30 mL/min/1.73m²) in LDKT. This integrative strategy has the potential to support posttransplant decision-making and improve transplant outcomes. Further validation in larger, multicenter cohorts is warranted to confirm the generalizability and clinical utility of this approach.

## Data Availability

All data produced in the present study are available upon reasonable request to the authors

## Notes

### Competing Interest Statement

The authors have declared no competing interest.

### Funding Statement

This study did not receive any funding

### Author Declarations

Ethics committee of Erasmus MC waived ethical approval for this work

