## Supplementary for "Machine Learning-Driven Assessment of Early Graft Function in Living Donor Kidney Transplantation Using Intraoperative Laser Speckle Contrast Imaging"

**1. Supplementary Methods**

**Methods S1: TRIPOD Checklist: Prediction Model Development and Validation**

| **Section/Topic** | **Item** |  | **Checklist Item** | **Page** |
| --- | --- | --- | --- | --- |
| **Title and abstract** | | | | |
| Title | 1 | D;V | Identify the study as developing and/or validating a multivariable prediction model, the target population, and the outcome to be predicted. | 1 |
| Abstract | 2 | D;V | Provide a summary of objectives, study design, setting, participants, sample size, predictors, outcome, statistical analysis, results, and conclusions. | 1, 2 |
| **Introduction** | | | | |
| Background and objectives | 3a | D;V | Explain the medical context (including whether diagnostic or prognostic) and rationale for developing or validating the multivariable prediction model, including references to existing models. | 3 |
|  | 3b | D;V | Specify the objectives, including whether the study describes the development or validation of the model or both. | 3, 4 |
| **Methods** | | | | |
| Source of data | 4a | D;V | Describe the study design or source of data (e.g., randomized trial, cohort, or registry data), separately for the development and validation data sets, if applicable. | 4 |
|  | 4b | D;V | Specify the key study dates, including start of accrual; end of accrual; and, if applicable, end of follow-up. | 4 |
| Participants | 5a | D;V | Specify key elements of the study setting (e.g., primary care, secondary care, general population) including number and location of centres. | 4 |
|  | 5b | D;V | Describe eligibility criteria for participants. | 4 |
|  | 5c | D;V | Give details of treatments received, if relevant. | NA |
| Outcome | 6a | D;V | Clearly define the outcome that is predicted by the prediction model, including how and when assessed. | 4 |
|  | 6b | D;V | Report any actions to blind assessment of the outcome to be predicted. | 4 |
| Predictors | 7a | D;V | Clearly define all predictors used in developing or validating the multivariable prediction model, including how and when they were measured. | 4-6 |
|  | 7b | D;V | Report any actions to blind assessment of predictors for the outcome and other predictors. | 9 |
| Sample size | 8 | D;V | Explain how the study size was arrived at. | 4 |
| Missing data | 9 | D;V | Describe how missing data were handled (e.g., complete-case analysis, single imputation, multiple imputation) with details of any imputation method. | NA |
| Statistical analysis methods | 10a | D | Describe how predictors were handled in the analyses. | 4-7 |
|  | 10b | D | Specify type of model, all model-building procedures (including any predictor selection), and method for internal validation. | 4-7 |
|  | 10c | V | For validation, describe how the predictions were calculated. | 7, 8 |
|  | 10d | D;V | Specify all measures used to assess model performance and, if relevant, to compare multiple models. | 7, 8 |
|  | 10e | V | Describe any model updating (e.g., recalibration) arising from the validation, if done. | NA |
| Risk groups | 11 | D;V | Provide details on how risk groups were created, if done. | NA |
| Development vs. validation | 12 | V | For validation, identify any differences from the development data in setting, eligibility criteria, outcome, and predictors. | 7 |
| **Results** | | | | |
| Participants | 13a | D;V | Describe the flow of participants through the study, including the number of participants with and without the outcome and, if applicable, a summary of the follow-up time. A diagram may be helpful. | 8 |
|  | 13b | D;V | Describe the characteristics of the participants (basic demographics, clinical features, available predictors), including the number of participants with missing data for predictors and outcome. | 8 |
|  | 13c | V | For validation, show a comparison with the development data of the distribution of important variables (demographics, predictors and outcome). | 8 |
| Model development | 14a | D | Specify the number of participants and outcome events in each analysis. | 8 |
|  | 14b | D | If done, report the unadjusted association between each candidate predictor and outcome. | 8 |
| Model specification | 15a | D | Present the full prediction model to allow predictions for individuals (i.e., all regression coefficients, and model intercept or baseline survival at a given time point). | 7, 25-28 |
|  | 15b | D | Explain how to the use the prediction model. | 25-28 |
| Model performance | 16 | D;V | Report performance measures (with CIs) for the prediction model. | 9 |
| Model-updating | 17 | V | If done, report the results from any model updating (i.e., model specification, model performance). | NA |
| **Discussion** | | | | |
| Limitations | 18 | D;V | Discuss any limitations of the study (such as nonrepresentative sample, few events per predictor, missing data). | 11 |
| Interpretation | 19a | V | For validation, discuss the results with reference to performance in the development data, and any other validation data. | 9-11 |
|  | 19b | D;V | Give an overall interpretation of the results, considering objectives, limitations, results from similar studies, and other relevant evidence. | 9-11 |
| Implications | 20 | D;V | Discuss the potential clinical use of the model and implications for future research. | 11, 12 |
| **Other information** | | | | |
| Supplementary information | 21 | D;V | Provide information about the availability of supplementary resources, such as study protocol, Web calculator, and data sets. | 22-33 |
| Funding | 22 | D;V | Give the source of funding and the role of the funders for the present study. | NA |

*Items relevant only to the development of a prediction model are denoted by D, items relating solely to a validation of a prediction model are denoted by V, and items relating to both are denoted D;V. We recommend using the TRIPOD Checklist in conjunction with the TRIPOD Explanation and Elaboration document.

**Methods S2: Laser Speckle Contrast Imaging (LSCI) Measurement Procedures**

**S2.1 Basic principle of LSCI**

When scattering tissue is illuminated with a coherent near-infrared laser, the scattered light interferes and produces a random speckle pattern, which is detected by a high-speed camera. The motion of red blood cells (RBCs) causes temporal fluctuations in this speckle pattern. Faster RBC motion results in faster speckle fluctuations and lower speckle contrast.

In spatial mode, a kernel size of 5×5 pixels is used to calculate the speckle contrast *K* using the equation:

$K=\frac{\sigma}{\left\langle I \right\rangle}$ (1),

where *σ* is the standard deviation of pixel intensity *I*, and $\left\langle I \right\rangle$ is the mean intensity within the kernel. In this mode, the basic formula for LSCI measurement of renal perfusion (Flux) is $Flux\propto\left( \frac{\left\langle I \right\rangle}{\sigma} \right)^{2}$ (2), and measured in perfusion unit (PU).

Higher RBC velocities lead to lower *K* and higher Flux values. As speckle patterns fluctuate rapidly with fast RBC motion, the camera's exposure time averages the signal, resulting in a reduced intensity variance (σ) and thus lower contrast.

**S2.2 LSCI Measurement Settings**

All LSCI measurements were conducted using the MoorO_2_Flo system (Moor Instruments Ltd., Axminster, UK), with a laser wavelength of 785 nm. The system was set to an exposure time of 20 ms and a spatial resolution of 3.9 μm per pixel. Measurements were performed peri-transplant during the reperfusion phase, ~15 minutes after vascular anastomosis, once perfusion of was stable. The kidney was positioned in its final transplant orientation. To eliminate external interference, ambient light was blocked by turning off surgical lights and closing window shades.

**S2.2.1 Measurement Distance**

As the distance between the camera and the sample increases, the speckle size increases. If the speckle size becomes larger than the camera pixel size, the speckle contrast artificially increases, leading to an overestimation of *K* and an underestimation of flux. To ensure consistency, a working distance of ~20 cm was maintained to keep the kidney fully in the field of view.

**S2.2.2 Camera Gain**

Higher camera gain leads to overestimation of RBC velocity, while lower gain leads to the loss of low velocity. A fixed gain of ~150 was used across all measurements to standardize results.

**S2.2.3 LED Brightness**

Higher brightness leads to overestimation of flux. Brighter illumination increases ⟨𝐼⟩ (mean intensity), therefore increases Flux. LED brightness was kept constant during all measurements.

**S.2.2.4 Measurement Frequency**

High frequency is suitable for faster-moving RBCs and low frequency for slower RBCs. As a frequency of 20 Hz is commonly used for low to moderate velocities, all measurements were performed at the rate of 20 Hz. Each measurement lasted for 10 seconds.

**S2.3 Quick Start Guide**

1. Make sure the system is turned on and that all leads are properly connected.

2. Open the **moorO2Flo Measurement** software.

3. Select the **Setup** tab and open the **Video Preview** window to position the camera to make sure the kidney is in the field of view.

4. Select the **Setup** tab and open the **System Setup** window to adjust the following settings:

- Adjust the **Zoom** setting to make sure the whole kidney is the field of view.
- Adjust the **Focus** setting to make sure the focus is on the subject. **Auto Focus** may be unsuccessful and manual adjustment may be required.
- Adjust the **Camera Gain** to 150.
- Adjust the **LED Brightness** to achieve an optimal brightness. For manual adjustment always set the camera gain first before adjusting LED brightness.

5. Select the **Image Settings** tab and configure the following settings:

- **High resolution** processing mode (576×748 pixels).
- **Frame rate**: **Frequency**, 20 Hz.
- Duration: **Fixed duration**, 10 s.

6. Close the System Setup window with the **OK** button.

7. Start the measurement by clicking **Start measurement** on the toolbar. Measurement should start immediately and the perfusion.

8. Once the set duration ends, the measurement will stop automatically. Click **Save As** to save the measurement data.

**S2.4 Real-Time Monitoring and Quality Control**

During LSCI acquisition, real-time renal perfusion was assessed. If abnormal perfusion patterns were detected, the transplant surgeons were notified to assess potential issues such as anastomotic stenosis or renal artery kinking. If any adjustments were made that may affect renal perfusion, another LSCI measurement would be performed included for further analysis.

**Methods S3: Description of the Machine Learning Classification Models**

**S3.1 Support Vector Machine**

Support Vector Classification is a supervised machine learning algorithm derived from classical Support Vector Machine (SVM) framework, designed to solve binary classification problems by finding the optimal separating hyperplane that maximizes the margin between two classes. The C-SVC formulation incorporates a regularization parameter **C**, which balances the trade-off between maximizing the margin and minimizing classification errors (soft-margin SVM).

In this study, linear kernel (LK-SVM) was applied.

The linear kernel computes the similarity between two input vectors $x_{i}$ and $x_{j}$ using a simple dot product: $K\left( x_{i},x_{j} \right)=x_{i}^{T}x_{j}$

This kernel assumes that the classes can be separated by a linear decision boundary. It does not require additional hyperparameters beyond the regularization parameter **C**, making it suitable for high-dimensional data or cases where the relationship between variables and outcome is approximately linear.

**S3.2 Classification Random Forest**

Classification Random Forest is a supervised machine learning method derived from the decision tree algorithm, designed to enhance prediction accuracy and reduce overfitting. It achieves this by constructing a large number of decision trees during training and aggregating their predictions to produce a final classification outcome.

Each individual tree in the forest is trained on a bootstrap sample, randomly drawn with replacement, from the original training dataset. To introduce further randomness and reduce correlation among trees, only a random subset of predictors is considered at each node when determining the best split.

Given a training set $D=\left\{ \left( x_{1},y_{1} \right),\left( x_{2},y_{2} \right),\ldots,\left( x_{n},y_{n} \right) \right\}$, where $x_{i}$ represents the feature vector and $y_{i}\in\left\{ 0, 1 \right\}$ is the class label (“Stable” = 0, “High-risk” = 1), the algorithm proceeds as follow:

For b = 1 to B (e.g., 500 trees)

- Bootstrap sample D_b_ is drawn from D.
- A decision tree T_b_ is trained on D_b_. m candidate predictors are selected at each node of the tree, and the split is made based on the Gini impurity.

For a new observation $x$, the forest predicts the class $\hat{y}$ by majority vote

- $\hat{y}=majority vote\left\{ T_{b}\left( x \right) \right\}_{b=1}^{B}$

**Split Rule**

At each node, the algorithm evaluates candidate splits using the Gini impurity criterion, defined as: $G\left( t \right)=1-\sum_{k=1}^{K} {p_{k}}^{2}$, where $G(t)$ is the Gini impurity of node $t$, $K$ is the number of classes (in binary classification, $K=2$), $p_{k}$ is the proportion of samples of class k in node t. The split that achieves the maximum reduction in impurity (i.e., the largest decrease in $G(t)$) is selected.

In this study, a Classification Random Forest was used to predict binary graft outcomes (“Stable” vs. “At-risk”) based on a combination of clinical and LSCI-derived features. The model was implemented using the “randomForest” package in R with the following parameters:

Number of trees (B) = 500

Features considered at each split $\left( m \right)=\sqrt{p}$, the default setting for classification.

**S3.3 Extreme Gradient Boosting**

Extreme Gradient Boosting (XGBoost) is a scalable and efficient implementation of gradient boosting machines, designed for supervised learning tasks including classification and regression. It builds an ensemble of decision trees sequentially, where each new tree is trained to correct the errors made by the previous ones. XGBoost improves upon traditional gradient boosting by introducing optimized regularization, parallel processing, and handling of missing values, which enhances both model performance and computational speed.

In XGBoost, the model minimizes a regularized objective function that combines a loss term $L(\phi)$ (e.g., logistic loss for binary classification) and a regularization term $\Omega(f)$ to control model complexity: $L(\phi)=\sum_{i=1}^{n} l\left( y_{i},\hat{y}_{i} \right)+\sum_{k=1}^{K} \Omega\left( f_{k} \right)$, where $\Omega\left( f \right)=\gamma T+\frac{1}{2}\lambda\sum_{j=1}^{T} {w_{j}}^{2}$,

$l\left( y_{i},\hat{y}_{i} \right)$ is the loss function (e.g., log-loss),

$f_{k}$ is the function (tree) added at boosting iteration $k$,

$T$ is the number of leaves in the tree,

$w_{j}$ are the leaf weights,

$\gamma$ and $\lambda$ are regularization parameters.

The model uses second-order gradient information (both first and second derivatives of the loss function) to guide tree construction and split evaluation, enabling more precise and efficient optimization.

**S3.4 Logistic Regression**

Logistic regression is a supervised machine learning algorithm commonly used for binary classification problems, where the outcome variable has two possible classes. It models the probability that a given input belongs to a particular class by applying the logistic (sigmoid) function to a linear combination of input features. The resulting output is a value between 0 and 1, interpreted as the probability of class membership.

In this study, we applied a binary logistic regression model to predict early graft dysfunction which required no hyperparameters. For classification, a default decision threshold of 0.5 was applied to the predicted probabilities. Cases with predicted probability ≥ 0.5 were classified as "At-risk", and those with probabilities < 0.5 were classified as "Stable".

**Table S1. Performance metrics of the ML models for early graft function classification in the oversampled train data**

| Modeling  Median (IQR) | Accuracy | Sensitivity  (Recall) | Specificity | Precision  (PPV) | NPV | F1 score | AUC  (ROC) |
| --- | --- | --- | --- | --- | --- | --- | --- |
| SVM | | | | | | | |
| Clinical | 0.79 (0.75-0.84) | 0.77 (0.73-0.80) | 0.83 (0.79-0.89) | 0.85 (0.80-0.91) | 0.83 (0.79-0.89) | 0.79 (0.76-0.85) | 0.84 (0.81-0.88) |
| Combined | 0.89 (0.85-0.92) | 0.88 (0.84-0.91) | 0.89 (0.88-0.95) | 0.89 (0.88-0.95) | 0.89 (0.88-0.95) | 0.89 (0.86-0.92) | 0.94 (0.91-0.96) |
| RF | | | | | | | |
| Clinical | 0.92 (0.90-0.92) | 0.89 (0.87-0.90) | 0.94 (0.91-0.97) | 0.94 (0.91-0.97) | 0.94 (0.91-0.97) | 0.92 (0.91-0.93) | 0.97 (0.96-0.98) |
| Combined | 0.95 (0.94-0.97) | 0.95 (0.94-0.97) | 0.96 (0.94-0.97) | 0.95 (0.94-0.97) | 0.96 (0.94-0.97) | 0.95 (0.94-0.97) | 0.99 (0.98-0.99) |
| XGBoost | | | | | | | |
| Clinical | 0.90 (0.88-0.92) | 0.88 (0.87-0.89) | 0.94 (0.90-0.95) | 0.94 (0.90-0.95) | 0.94 (0.90-0.95) | 0.90 (0.88-0.92) | 0.96 (0.95-0.97) |
| Combined | 0.94 (0.92-0.95) | 0.93 (0.90-0.94) | 0.95 (0.94-0.97) | 0.95 (0.94-0.97) | 0.95 (0.94-0.97) | 0.94 (0.92-0.95) | 0.98 (0.98-0.99) |
| LR | | | | | | | |
| Clinical | 0.79 (0.77-0.81) | 0.76 (0.74-0.79) | 0.82 (0.79-0.86) | 0.83 (0.81-0.86) | 0.82 (0.79-0.86) | 0.80 (0.77-0.82) | 0.84 (0.82-0.87) |
| Combined | 0.88 (0.85-0.91) | 0.86 (0.83-0.88) | 0.89 (0.87-0.95) | 0.89 (0.87-0.95) | 0.89 (0.87-0.95) | 0.88 (0.85-0.91) | 0.92 (0.91-0.94) |

AUC, area under the curve; IQR, interquartile range; ML, machine learning; NPV, negative predictive value; PPV, positive predictive value; RF, random forest; ROC, receiver operating characteristic; SVM, support vector machine; XGBoost, extreme gradient boosting.

**Table S2. Performance metrics of the ML models for early graft function classification in the test data**

| Modeling  Median (IQR) | Accuracy | Sensitivity  (Recall) | Specificity | Precision  (PPV) | NPV | F1 score | AUC  (ROC) |
| --- | --- | --- | --- | --- | --- | --- | --- |
| SVM | | | | | | | |
| Clinical | 0.77 (0.72-0.80) | 0.29 (0.21-0.37) | 0.95 (0.91-0.96) | 0.75 (0.50-0.75) | 0.95 (0.91-0.96) | 0.43 (0.31-0.53) | 0.79 (0.72-0.89) |
| Combined | 0.88 (0.84-0.91) | 0.50 (0.41-0.60) | 0.96 (0.93-0.96) | 0.75 (0.50-0.75) | 0.96 (0.93-0.96) | 0.60 (0.50-0.71) | 0.84 (0.81-0.95) |
| RF | | | | | | | |
| Clinical | 0.84 (0.81-0.88) | 0.33 (0.25-0.50) | 0.90 (0.89-0.93) | 0.25 (0.25-0.50) | 0.90 (0.89-0.93) | 0.40 (0.29-0.50) | 0.77 (0.72-0.85) |
| Combined | 0.91 (0.88-0.91) | 0.88 (0.50-1.00) | 0.90 (0.90-0.93) | 0.25 (0.25-0.50) | 0.90 (0.90-0.93) | 0.40 (0.35-0.64) | 0.90 (0.77-0.96) |
| XGBoost | | | | | | | |
| Clinical | 0.81 (0.78-0.87) | 0.25 (0.14-0.40) | 0.89 (0.88-0.92) | 0.25 (0.25-0.50) | 0.89 (0.88-0.92) | 0.36 (0.22-0.45) | 0.76 (0.66-0.83) |
| Combined | 0.88 (0.84-0.91) | 0.50 (0.33-0.67) | 0.90 (0.90-0.95) | 0.25 (0.25-0.69) | 0.90 (0.90-0.95) | 0.42 (0.33-0.65) | 0.84 (0.72-0.88) |
| LR | | | | | | | |
| Clinical | 0.75 (0.72-0.78) | 0.29 (0.23-0.33) | 0.93 (0.91-0.96) | 0.50 (0.50-0.75) | 0.93 (0.91-0.96) | 0.42 (0.33-0.46) | 0.78 (0.69-0.88) |
| Combined | 0.88 (0.81-0.91) | 0.50 (0.38-0.59) | 0.96 (0.93-0.96) | 0.75 (0.50-0.75) | 0.96 (0.93-0.96) | 0.60 (0.49-0.67) | 0.82 (0.79-0.92) |

AUC, area under the curve; IQR, interquartile range; ML, machine learning; NPV, negative predictive value; PPV, positive predictive value; RF, random forest; ROC, receiver operating characteristic; SVM, support vector machine; XGBoost, extreme gradient boosting.

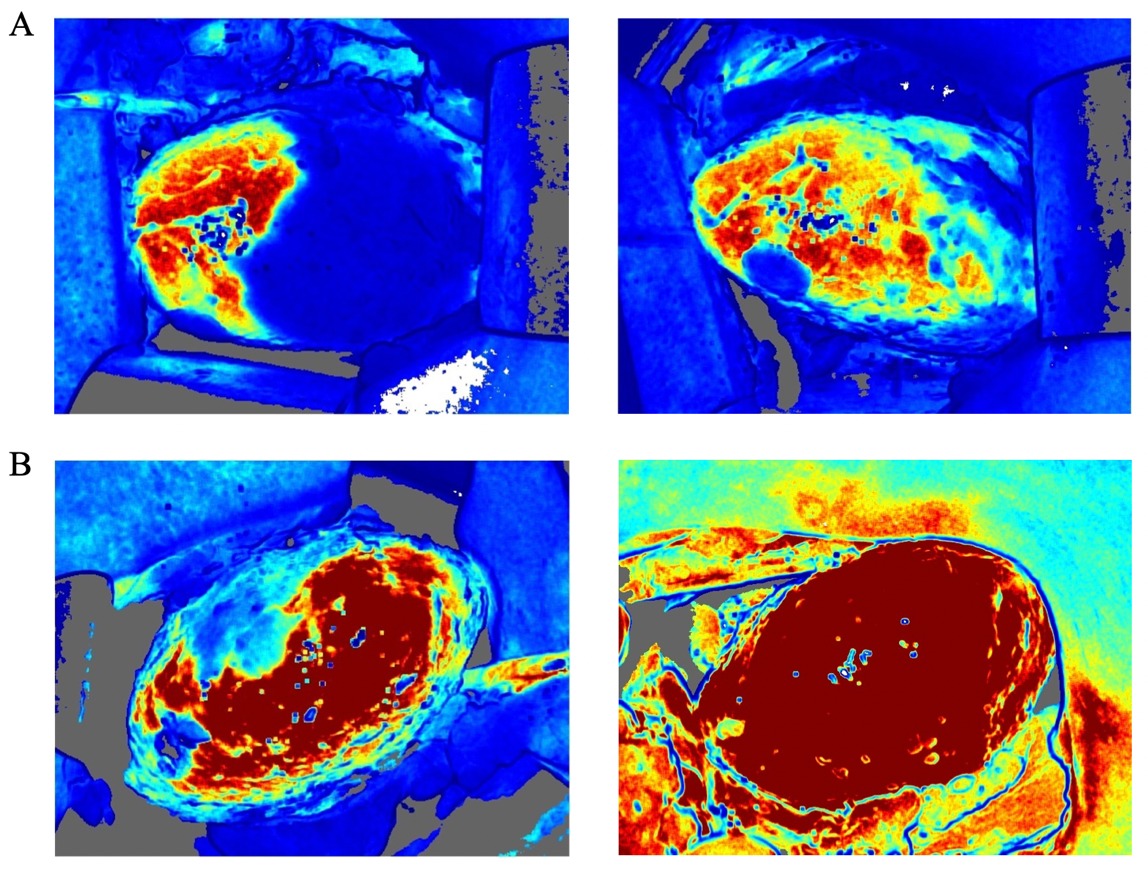

**Figure S1.** Representative LSCI images of two kidney transplant cases showing intraoperative perfusion abnormalities and improved renal cortical perfusion following intervention. (A) The kidney required arterial reconstruction due to the presence of two donor renal artery branches without a patch. LSCI revealed extensive cortical perfusion deficiency in the upper pole after initial reperfusion (left). The arterial anastomosis was removed and reconstructed, and subsequent LSCI demonstrated improved perfusion (right); (B) LSCI revealed a localized perfusion defect after reperfusion (left). Intraoperative assessment identified renal artery kinking, which was corrected by adjusting the kidney’s position. Subsequent LSCI imaging confirmed restoration of adequate perfusion (right).

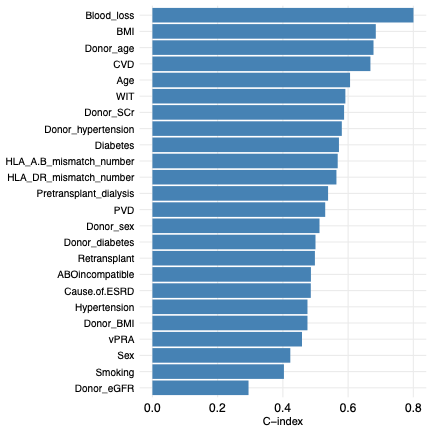

**Figure S2.** Concordance index (C-index) of all candidate predictor variables used for Clinical Model development. Each bar represents the C-index of a single variable in discriminating between “Stable” and “High-risk” graft outcomes.

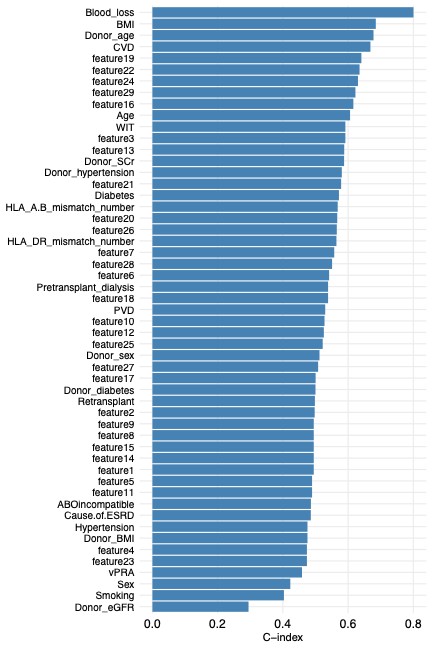

**Figure S3.** Concordance index (C-index) of all candidate predictor variables used for Combined Model development. Each bar represents the C-index of a single variable in discriminating between “Stable” and “High-risk” graft outcomes.
